# Risk factors for bacterial infections in patients with moderate to severe COVID-19: A case control study

**DOI:** 10.1101/2021.01.09.21249498

**Authors:** N. Nasir, F. Rehman, S.F. Omair

**Affiliations:** Dept. of Medicine, Aga Khan University, Karachi, Pakistan

**Keywords:** Bacteria, COVID-19, nosocomial infections, co-infection

## Abstract

**Objective:** Bacterial infections are known to complicate respiratory viral infections and are associated with adverse outcomes in COVID-19 patients. A case control study was conducted to determine risk factors for bacterial infections where cases were defined as moderate to severe/critical COVID-19 patients with bacterial infection and those without were included as controls. Logistic regression analysis was performed.

**Results:** Out of a total of 50 cases and 50 controls, greater proportion of cases had severe or critical disease at presentation as compared to control i.e 80% vs 30% (p<0.001). Hospital acquired pneumonia (72%) and Gram negative organisms (82%) were predominant. Overall antibiotic utilization was 82% and was 64% in patients who had no evidence of bacterial infection. The median length of stay was significantly longer among cases compared to controls (12.5 versus 7.5 days) (p=0.001). The overall mortality was 30%, with comparatively higher proportion of deaths among cases (42% versus 18%) (p=0.009). Severe or critical COVID-19 at presentation (AOR: 4.42 times; 95% CI; 1.63-11.9) and use of steroids (AOR: 4.60; 95% CI 1.24-17.05) were independently associated with risk of bacterial infections. These findings have implications for antibiotic stewardship as antibiotics can be reserved for those at higher risk for bacterial superinfections.

## INTRODUCTION

COVID-19 pandemic has claimed more than 1,000,000 lives to date and its long term impact is yet to be determined. Respiratory viral infections have been well known to predispose patients to co-infections and these lead to increased disease severity and mortality as was observed in 1918 influenza outbreak, where most mortalities were due to simultaneous bacterial infection.[1] Bacterial co-infection also led to poor outcome in 2009H1N1 influenza pandemic.[2]

The incidence of bacterial co-infection in COVID-19 ranges from 3-30% [3, 4]. Zhou and colleagues showed that in the current coronavirus disease 2019 (COVID-19) pandemic, 50% of patients who died, had secondary bacterial infections, while another study showed presence of both bacterial and fungal infection [5, 6]. Due to similar clinical phenotype and difficulties in identifying COVID- 19 disease from atypical bacterial pneumonia or nosocomial pneumonia some guidelines advise empirical antibiotics [7]. In a study conducted at 38 hospitals in Michigan 56.6% patients received empirical antibiotics therapy[8].

Studies indicate that bacterial co-infection and secondary infection complicate COVID-19 though regional data is scarce[6]. Moreoverrisk factors for these infections need to be better elucidated. Antimicrobials are used empirically which may lead to antimicrobial resistance in the long term. Hence, it is imperative to conduct comparative studies to identify those COVID-19 patients who are candidates for empirical antibiotic therapy and curtail widespread injudicious use of antibiotics.The purpose of this study is to determine the risk factors of bacterial infections in patients and characterization of these infections along with the sensitivity patterns of isolated organisms in patients presenting with moderate to severe/critical COVID-19.

## MAIN TEXT

### METHODS

A case control study was conducted from February 2020 to June 2020 at a tertiary care center in Karachi, Pakistan. Cases were defined as patients who had PCR confirmed moderate to severe/critical COVID-19 as per WHO criteria for severity and had evidence of bacterial infection based on isolation of bacteria in any of the culture specimens collected during admission along with symptoms and signs consistent with infection. Controls were defined as patients who had PCR confirmed moderate to severe/critical COVID-19 as per WHO criteria for severity but who did not develop bacterial infection during admission.

All adult patients (age >=18 years) hospitalized with moderate and severe/critical COVID-19 as per WHO definition for severity [9] at Aga Khan University Hospital, Karachi were assessed for presence of bacterial co-infection at admission as well as followed for secondary bacterial infection during hospitalization. Patients with signs and symptoms consistent with bacterial infection during hospitalization with COVID-19 at various sites such as urinary tract infection, hospital acquired pneumonia, ventilator associated pneumonia, central line associated blood- stream infection etc. as defined by CDC [10] and who were identified to have growth of significant bacteria in the respective appropriate culture specimens (e.g urine, tracheal aspirate, blood from central line etc) collected during admission were included whereas those patients who had evidence of bacterial infection on culture but did not have signs and symptoms consistent with infection at any site as defined by CDC [10] were considered to be colonized and excluded from study and patients with mono-microbial fungal infections were also excluded.

Patients who had any positive bacterial culture were identified from infection control records and were screened for eligibility criteria for cases. Controls were identified from medical records among all moderate to severe/critical COVID-19 admissions. A sample size of 50 cases was obtained. For each case patient, 1:1 control patient(s) was also obtained (50 controls). Main outcome variable was presence of bacterial infection defined as infection with clinically significant bacteria which is identified from a culture specimen among those with moderate or severe/critical COVID-19. Exposure variables included patient related factors such as age in years, gender, comorbidities, type of ward/unit to which patient was admitted, presence of invasive devices and immunosuppression received. We collected data on potential confounders including previous co-morbids and severity of illness. Data was collected on structured proforma which was pretested for one week at the start of the study. Patients were further stratified as having community-onset infection if they had culture specimen positive within 72 hours of admission and hospital-onset infection if they were found to have a bacterial infection after 72 hours of hospitalization. Bacteriological identification was performed in College of American Pathologists (CAP) certified Clinical Microbiology laboratory at Aga Khan University by conventional methods. Antibiotic susceptibility of isolated bacteria was evaluated by the standard disc diffusion method in accordance with the Clinical & Laboratory Standards Institute (CLSI) recommendations.

#### Statistical analysis

Frequencies with percentages were reported for categorical variables such as gender, type of infection by site, etc. according to case or control status. For continuous variables such as age, length of stay etc. median and interquartile range were reported by case and control status. Logistic regression analysis was performed to determine the association between risk factors and bacterial infection in moderate and severe/critical COVID-19 patients and the results were reported as adjusted odds ratios with 95% confidence intervals. P-value 0.05 was considered significant. Data was analyzed using Stata version 12.

The study received an exemption from ethical approval from the Aga Khan University Ethics Review Committee (ERC reference number: 2020-5178-14123).

## RESULTS

A total of 50 cases and 50 controls were included in the analysis after making relevant exclusions over a period of 4 months from February to June 2020. Median age of cases was 58 years (IQR) and of controls was 62 years (IQR). The overall male to female ratio was 2.8 and gender distribution was similar between the groups. The most frequent co-morbids were diabetes and hypertension in both groups. The severity of illness was significantly different in both the groups with a high proportion (80%) of severe/critical COVID-19 patients among cases compared to 30% in controls (p<0.001). Among those COVID-19 patients with bacterial infections, the majority (72%) was hospital-acquired and 28% were community-acquired at onset and the commonest site of infection was hospital-acquired pneumonia (Figure 1). Nine out of 50 had bacteremia. Gram-negative infections were more common (82%) than gram-positive infections. The most frequently isolated organism from sputum specimens was multi-drug resistant Acinetobacter species in 27%, followed by multi-drug resistant *Pseudomonas aeroginosa* in 25% (Figure 2). Among patients with community-acquired infections, the most common organism was *Staphylococcus aureus* (5/14) in which four were methicillin-resistant and extended spectrum beta lactamase producing (ESBL) *Klebsiella pneumoniae* in (9/14). Majority of patients had a mono-microbial infection (70%). Seven out of 50 patients had simultaneous infection at two sites.

**Figure 1:**
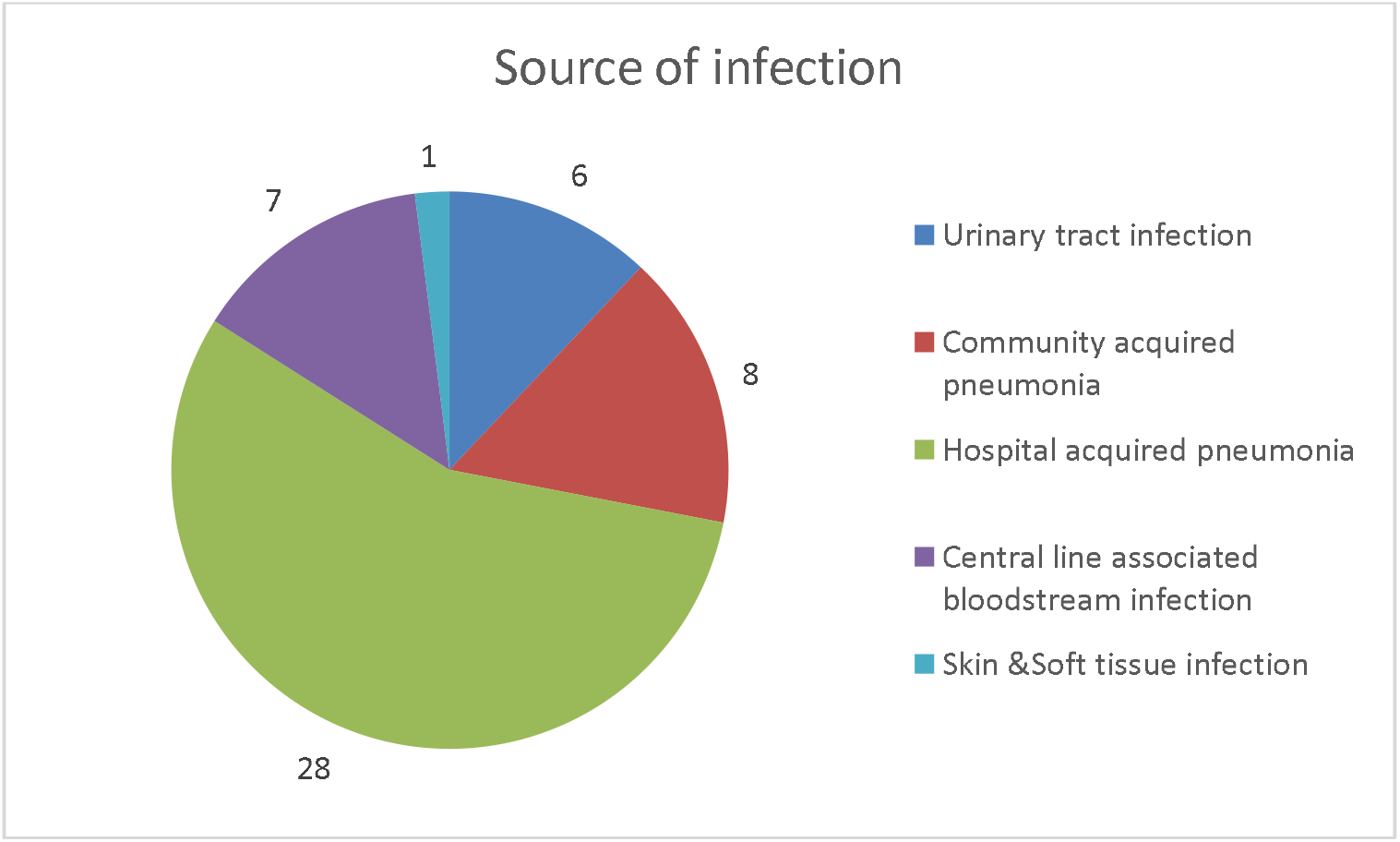
Sites of Infection in moderate and severe COVID-19 patients with bacterial infections.

**Figure 2:**
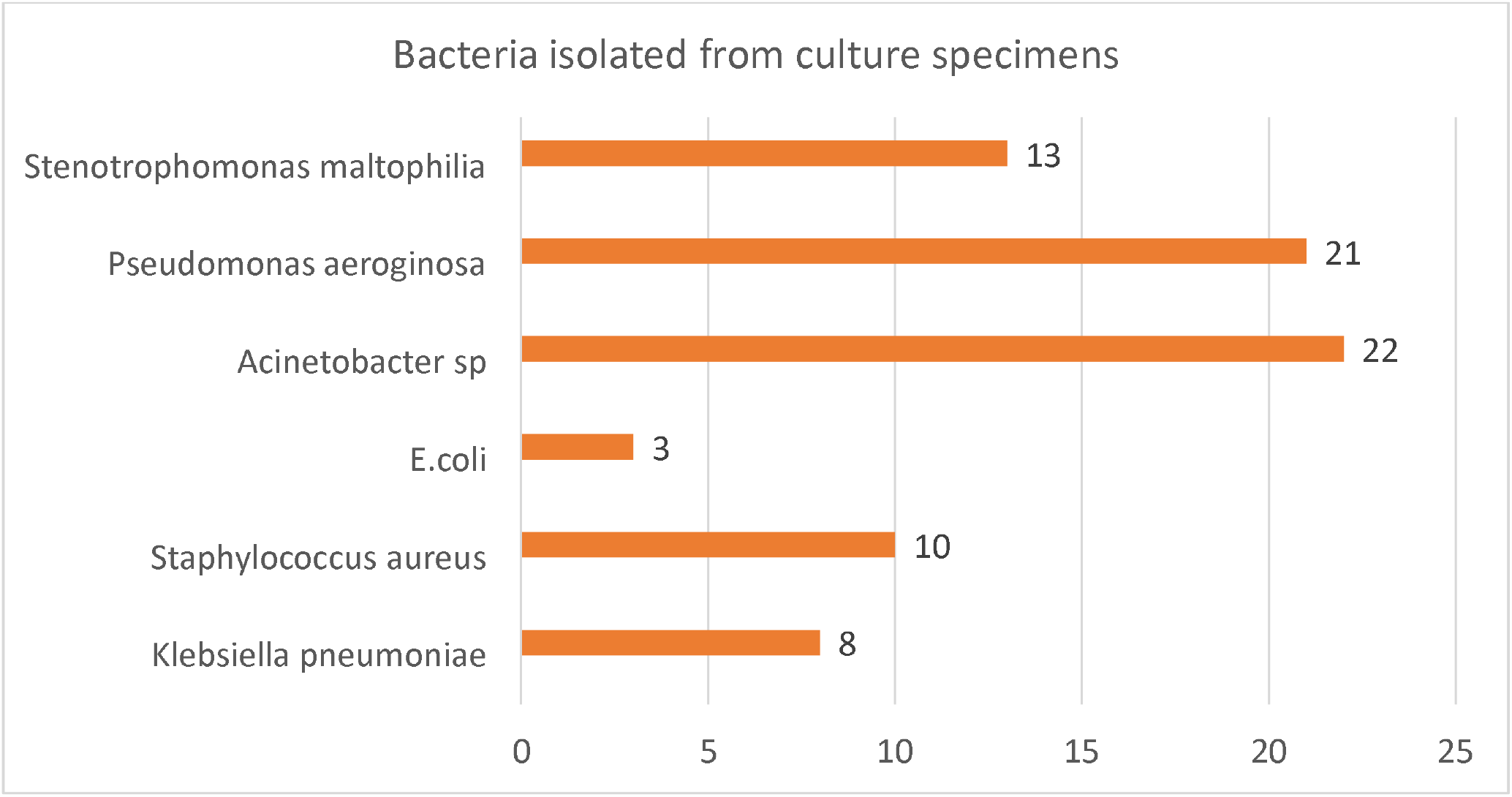
Clinically significant isolates from culture specimens in patients with moderate and severe COVID-19.

Among laboratory investigations, the median C-reactive protein and median neutrophil to lymphocyte ratio were significantly higher in cases compared to controls (Table 1). However, there was no statistically significant difference between procalcitonin levels in COVID-19 patients with bacterial infection compared to those without bacterial infection (p=0.883). With regard to the unit of admission, COVID-19 patients with bacterial infections were more frequently admitted to the Intensive care unit (56%) compared to patients without bacterial infections who were mostly admitted to the ward (38%) (p<0.001). The use of invasive devices such as endotracheal tube and central venous catheters were also more frequent among cases compared to controls (p<0.001) (Table 1). Patients with bacterial infections were managed with invasive ventilation in 56% of cases compared to 14% controls (p<0.001) and with non-invasive ventilation in 64% of cases compared to 34% controls (p=0.003). Comparatively higher proportion of patients who had bacterial infections had received treatment with systemic steroids (92%) (p=0.001). All patients with bacterial infections had received antibiotics and among controls (32/50) had received antibiotics. The choice of empiric antibiotics was based on local antibiogram and institutional guidelines for community acquired pneumonia and definitive antibiotic treatment was decided based on identification and sensitivity pattern of the isolated organism. The overall mortality was 30%. Patients with COVID-19 having bacterial infections had a comparatively greater proportion of deaths (42% versus 18%) (p=0.009). Out of 21 patients who died, 18 had infection with gram negative organism with Acinetobacter sp (n=9) and *Pseudomonas aeroginosa* (n=7) being most common isolates. The median length of stay was also significantly longer among cases compared to controls (12.5 versus 7.5 days) (p=0.001). Multivariable logistic regression showed that patients who were severe to critically ill at the time of admission with COVID-19 were 4.42 times (95% CI; 1.63-11.9) at risk for bacterial infection and treatment with steroids was also a significant risk factor (AOR: 4.60; 95% CI 1.24-17.05). Admission to ward unit was found to be protective (OR: 0.15; 95% CI 0.02-0.75).

**Table 1:**
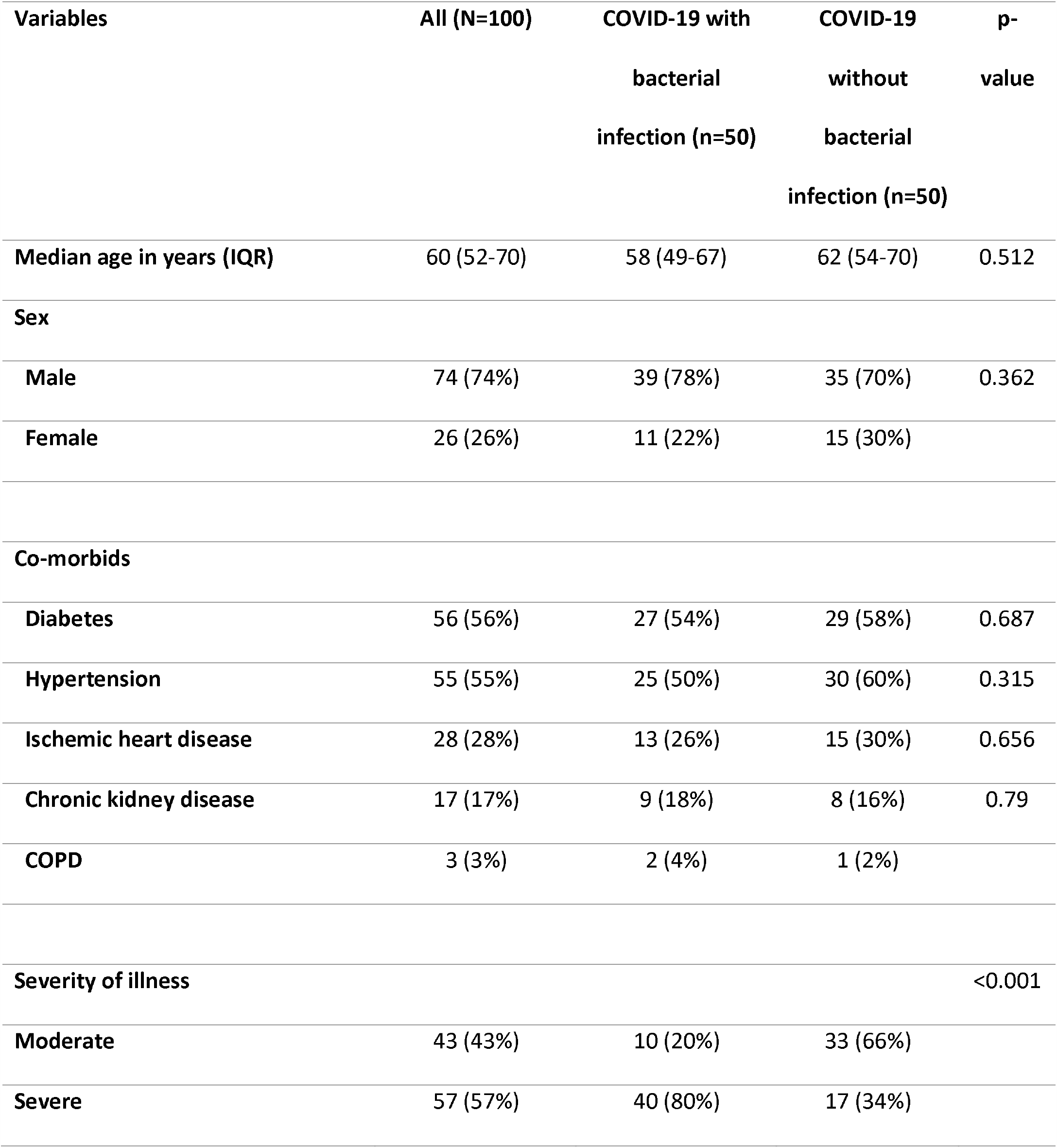
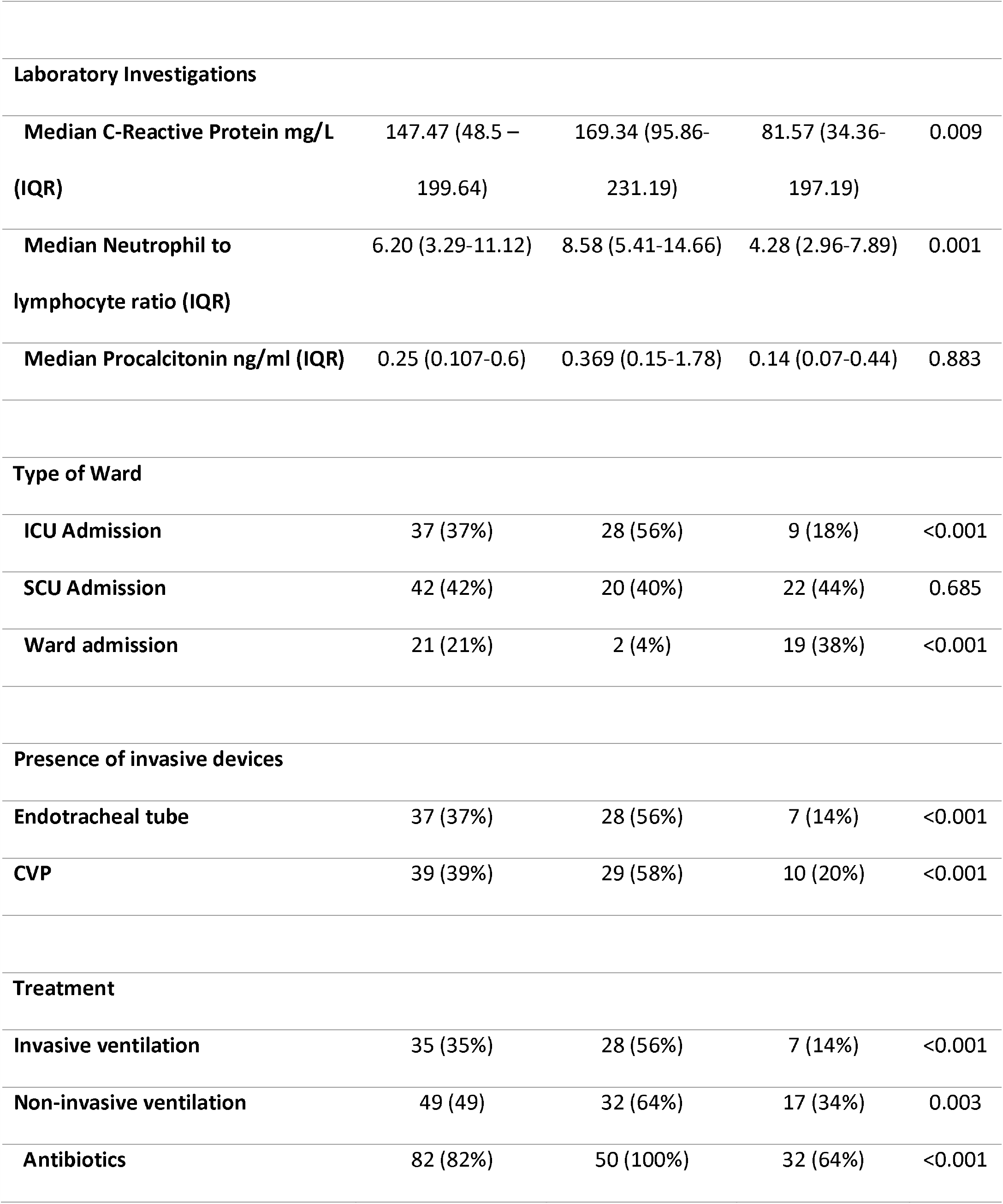
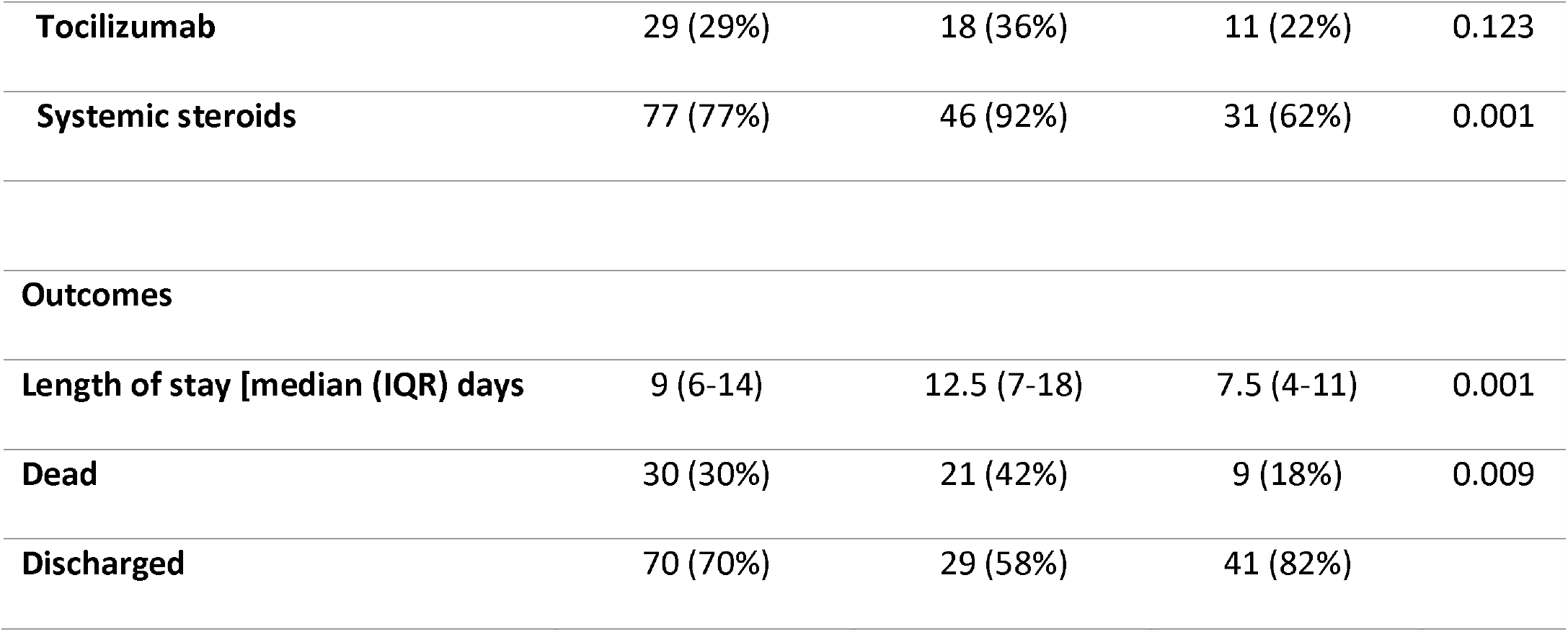
Comparison of moderate and severe COVID-19 patients with and without bacterial infections

## DISCUSSION

Our study found that there is a significant association of severity of illness with COVID-19 at presentation and treatment with steroids with the development of superimposed bacterial infection. Our patient cohort had moderate to severe or critical COVID-19 at the time of hospitalization and we had excluded patients who had specimen positive with contaminants or who were presumed to be colonized and without clinically significant infection. There was a high male to the female ratio and several studies have reported a greater risk of severe illness with COVID-19 in males compared to females [11, 12]. We had very few community-acquired infections and the majority were hospital-acquired. This is similar to study conducted by Hughes et al. from UK where only 3.2% co-infections were reported [13] and a study from Spain describing the incidence of co-infections [14]. This was also confirmed in a meta-analysis where the reported prevalence of co-infections was 3.5% [15]. Majority of the patients in our cohort developed hospital acquired infections. This is consistent with other reported literature [15, 16]. The most common site of infection was pneumonia and this is in concordance with initial reports from China [16]. Gram-negative infections have dominated as far as the type of organisms is concerned and this is similarly seen in studies reported from other parts of the world describing superinfections or secondary bacterial infections [14, 16]. There is a lack of reported literature on the sensitivity patterns of organisms isolated. In our study, we found multi-drug resistant Acinetobacter as the predominant pathogen causing hospital acquired infections and methicillin resistant *Staphylococcus aureus* as the main cause of co-infection in COVID-19 in our patients. A case series of 19 patients from Iran has described *Acinetobacter baumanii* in majority of patients requiring ICU admission[17] which reflects epidemiological similarity to Pakistan. In our study, the overall mortality was 30% and was higher in those who had bacterial infections. This is also consistent with data reported from Europe and parts of Asia [15, 18]. Moreover, we found that overall antibiotic utilization was 82% and was 64% in patients who had no evidence of bacterial infection. This has also been similarly reported [18] indicating widespread injudicious use of antibiotics which can potentiate the problem of antimicrobial resistance. In our study, we did not find procalcitonin to be a reliable marker of distinguishing patients who had bacterial infections and those who did not have bacterial infections. Although studies have described procalcitonin levels to remain normal in severe COVID-19, whether they increase in bacterial infections has not been reported and indicates need of large comparative studies exploring this relationship[19]. Our study is the first comparative study with regard to bacterial infections in the setting of moderate and severe to critical COVID-19 and describes the sensitivity pattern of organisms in addition to clearly classifying them into community and hospital acquired infections.

## Conclusions

Our study highlights need for improving antibiotic stewardship practices and reserving antibiotics for those who are severely ill with COVID-19 at presentation and require treatment with systemic steroids and provides insight into the lack of utility of serum procalcitonin as a marker of bacterial sepsis in this setting.

## LIMITATIONS

Our study is limited as it’s a single center experience which might affect generalizability. Moreover, we did not have molecular methods and genotyping available for our bacterial isolates and information on the isolation rate of the prevalent Gram negatives and MRSA in non-COVID-19 patients over the study period was not available to draw comparisons with rates of infection seen in COVID-19 patients.

## Data Availability

All data generated or analysed during this study are included in this published article

## List of Abbreviations

IQR: Interquartile range
ERC: Ethics review committee
ESBL: Extended-spectrum Beta –lactamase
C-RP: C-reactive protein
COVID-19: Coronavirus infection disease-19
MDR: Multi-drug resistant
MRSA: Methicillin-resistant Staphylococcus aureus

## DECLARATIONS

### Ethics approval and consent to participate

The study was submitted for ethical approval to the Aga Khan University Ethics review committee and received exemption (ERC reference number: 2020-5178-14123). Consent to participate was not required because there was no direct interaction with participants. The study was eligible for exemption as data was collected from records and no personal identifiers were recorded or used.

### Consent for publication

Not applicable

### Availability of data and materials

All data generated or analysed during this study are included in this published article

### Authors’ contributions

NN conceived idea, major contributer to manuscript, collected and analyzed data

FR conceived idea, contributed to manuscript, collected data

FU collected data and contributer to manuscript

All authors read and approved the manuscript

## Acknowledgements

Not applicable

